# cliexa-RA Implementation in Colorado Arthritis Center: A Case Study of Quadruple Aim Impacts

**DOI:** 10.64898/2026.03.29.26349644

**Authors:** Ashley Darnell, Robert Spencer, David Silverman

## Abstract

**Background:** Research supporting the use of digital platforms to increase efficiency in clinical settings has emerged and yet implementation remains a challenge. This can be explained by the unique needs of clinics for data collection and electronic medical record integration.

**Objective:** To identify how screening and monitoring of Rheumatoid Arthritis patients through a customized electronic platform, cliexa-RA impacts patient experience, physician experience, cost of care, and population health based on the Institute for Healthcare Improvement’s quadruple aim.

**Methods:** cliexa-RA was delivered on three tablets at the Colorado Arthritis Center over a six-month period to patients and physicians, who were asked to complete a 16-question intake form allowing patients to score their ability to complete daily tasks using the RAPID3 scoring system, and a six-question patient engagement survey. The physician would then input 28 joint assessment scores following a physical examination. cliexa-RA would then calculate five disease state scores, DAS28 (ESR), DAS28 (CRP), SDAI, CDAI, RAPID3, and send an EMR-compatible PDF file.

**Results:** Time stamp and patient satisfaction data was collected on 300 patients. Patient intake forms and self-reporting took an average of 2.4 minutes, and clinic-reported time required for calculation and transcription of the data using cliexa-RA was 1 minute with an additional 10 second direct data integration to the EMR after form submission. Eighty-five percent of patients said they would recommend cliexa forms to other clinicians. cliexa-RA scored an average of 3.57 out of 4 when compared to paper in ease of use, 3.61 in patient-reported reduction of time spent, and 3.50 when asked how easy the platform was to understand. Overall patient satisfaction was scored at 3.55 out of 4 and physician experience was measured by the adoption of the program in the study clinic with full integration into the Greenway Health EMR (currently integration is pending). Cost of care and population health impacts were not immediately available as a result of the pilot study; however, numerous savings and improvement opportunities exist as a result of cliexa-RA implementation.

**Conclusion:** Patient experience and physician engagement had immediate positive impacts from the implementation of cliexa-RA. Cost of care opportunities exist in both time savings and annual Medicare reporting costs. Population health opportunities exist in the collection of patient data, increasing informed decision making by physicians, as well as in the potential for future RA research using this data.

## Introduction

### Background

Nearly 1.5 million people in the United States suffer from Rheumatoid Arthritis (RA) (1), an autoimmune disease that causes the body to attack the joints. If untreated, RA can cause permanent joint damage or deformity, prompting physicians to seek early, aggressive treatment. Treatment involves a complex combination of lifestyle changes and a wide range of pharmacologic treatments including corticosteriods, synthetic disease modifying anti-rheumatic drugs (sDMARDs), targeted synthetic DMARDs (tsDMARDs), and biologic DMARDs (bDMARDs) (2), each with varying degrees of effectiveness and risk. Currently, rheumatologists address the complexity of treatment through face-to-face appointments where patients spend approximately 84 minutes in the clinic filling out paperwork, waiting for the physician, and in the exam room, with an average wage loss of $43 per appointment (3). Physicians’ typical non-complex payment code assumes that the physician will spend roughly 15 minutes with the patient (4), 37% of that time was found to be dedicated to electronic health recording and desk work (5).

Rheumatoid arthritis treatment differs from many other chronic diseases in that there is not one common “gold standard” biomarker to base clinical decisions on. Instead, treatment determinations are made using a scoring process assessing both patient self-reported symptoms and a physical examination. One study found that the patient history component was deemed as the most important factor in diagnosing and managing RA for 64% and 74% of rheumatologists, respectively (1). Scores are hand calculated by physicians and due to time limitations, are often limited to a single set of disease activity scores. These scores are not always consistently stored in the patient’s EMR, hindering the ability to track disease progression over time, and patients have little access to their score results or trends. Common themes among physicians suggest that a consistent and accurate patient history is critical to identifying RA flare patterns and determining the proper dosing for treatments to manage pain and limit side effects (6). Despite this, rheumatologists typically do not identify mobile tools as a valid solution to this issue because of the “lack of relevant data capture,” (7).

The Colorado Arthritis Center (CAC), P.C., is a rheumatology clinic in Englewood, Colorado that specializes in the diagnosis and treatment of autoimmune and inflammatory diseases, (8). The clinic traditionally utilized a paper-form intake process, which required hand calculations that many physicians weren’t able to complete due to time constraints. This gap was seen in not only the data output provided for patients but also in the organization of how information was stored. Physicians also face challenges completing the requirements for the Centers for Medicaid and Medicare Services (CMS) merit-based incentive payment system (MIPS), a 9-point scoring system to ensure quality assurance for which CAC must submit specific data for reimbursement (personal communication by Robert Spencer, MD, 2018). To address these challenges, the Colorado Arthritis Center partnered with cliexa^®^ to conduct the pilot study for the cliexa-RA platform.

### Previous Work

cliexa-RA was developed by cliexa^®^ as part of the product suite “enabling chronic disease patients to track their disease using intuitive graphics, clinically validated scoring models and real-time data feeds to EMR systems.” (10). Cliexa-RA was ranked top 19 in the 2017 JMIR mHealth publication *Apps to People with Rheumatoid Arthritis to Monitor Their Disease Activity: A Review of Apps for Best Practice and Quality*. Among the top ranked, cliexa-RA was the only platform that had validated disease activity calculations, data tracking tools, 28 tender and swollen joint counts, and remote patient-care team engagement (1). The application uses a RAPID3 scoring system to analyze both patient self-reporting and physician physical examination data to create a 5-score report that can be directly uploaded to the patient’s EMR. Patients are able to download the application to their mobile device and self-report symptoms, medication adherence, and track pain over time.

### Impact Assessment Framework

The Institute for Healthcare Improvement (IHI) originally developed the *Triple Aim* framework for healthcare improvement, measuring patient experience, the cost of care and population health (11). Since then, the IHI has supported the implementation of an additional measure, physician experience, creating the *quadruple aim* (12). (See Figure 1).

**Figure 1.**
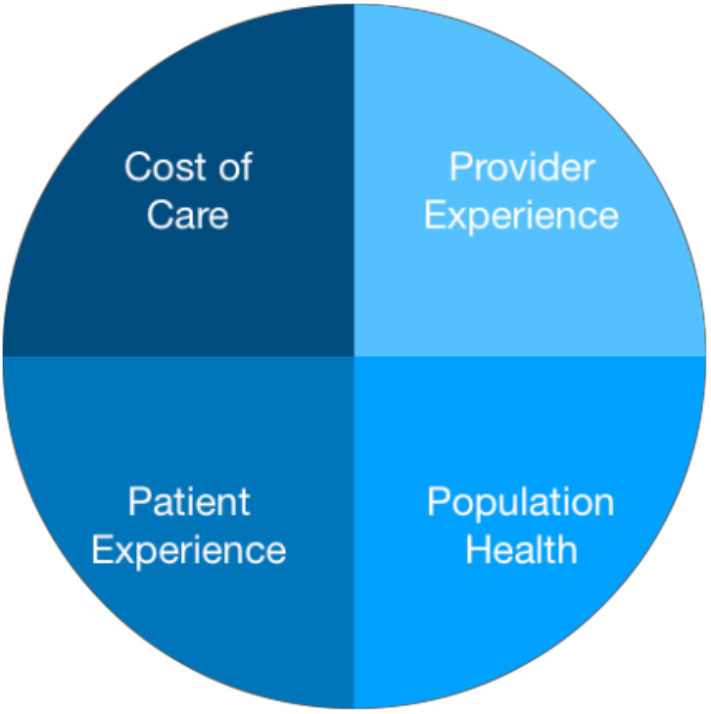
The Quadruple Aim Framework.

According to the CDC, national costs associated with arthritis were $140 billion in 2013, with nearly half owing to ambulatory care (13). Due to payer limitations and physician capacity, RA follow up appointments are typically scheduled every two months. This limits the availability for immediate follow up with physicians regarding symptoms caused by treatment transitions. Lag time between office visits without opportunity for follow up may contribute to the prevalence of ER visits and hospitalizations. The potential reduction in ER visits and hospitalizations could contribute to cost savings at the healthcare level,and increase patient satisfaction. Utilizing a digital tool in a clinic setting may increase consistency and uniformity in data collection and reporting, saving physicians time as well as meeting national quality assurance reporting measures.

The goal of this study is to assess quality impacts through the lens of the *Quadruple Aim* during the six-month pilot study implementing the digital tool, cliexa-RA, in the Colorado Arthritis Center.

## Methods and Data

The cliexa team worked with one of the authors, a rheumatologist, to create the patient intake form with Colorado Arthritis Center’s existing questionnaire using the RAPID3 assessment, a collective score based on the three patient-reported data measures from the American College of Rheumatology: function, pain, and patient global estimate of status (14). They also developed a patient engagement survey, and customized an additional step allowing clinicians to add physical exam data to allow calculation of disease activity scores. The output data later was expanded to include five scores, DAS28 (ESR), DAS 28 (CRP), SDAI, CDAI, and RAPID3. A RAPID3 score was calculated for each patient, the remaining scores were based upon the extent of physician input.

## Data Collection

The cliexa-RA digital platform was loaded onto tablets with Android and iOS capabilities. Three tablets were distributed to the Colorado Arthritis Center with cliexa-RA installed with customized assessment models. Devices were restricted to only utilize the cliexa-RA platform and were remotely managed by cliexa’s engineering team for troubleshooting. Data was gathered using DAS28, RAPID3, and a custom assessment questionnaire used by the Colorado Arthritis Center care team. The platform was piloted for 6 months and data was gathered on 300 patients through time stamp data collected by the platform and digital surveys.

### Process of Assessment

Patients arriving for their CAC appointments checked in with medical staff who entered the patient’s ID number into the platform which was used as an identifier in the EMR and restricted cliexa-RA from collecting patient identification data. The patient was given the tablet and completed the 16-question assessment and received a 6-question patient engagement survey to provide feedback on the cliexa-RA process. Once submitted, a cliexa message directed the patient to hand off the tablet to medical staff in the exam room. The clinician then opened a screen with fields that are specific to physical examination of the patient. After the examination was complete and scores were entered, cliexa-RA recalculated assessment scores and generated a PDF which was automatically emailed to the clinician. The clinician could then review the report, and download and attach the document to patients EMR record. The patient’s time to submit digital forms using cliexa-RA and the patient engagement survey responses were collected automatically in cliexa servers during the case study. Data management and analysis was conducted using Microsoft Excel.

## Results

Results are based on self-reporting of clinic staff, time stamp data, and survey results for 300 patients. Using cliexa-RA MOD, patient intake forms and self-reporting took an average of 2.4 minutes (see Figure 2), as compared to the standard estimate for paper form completion of 5 minutes (14), a 52% reduction. Clinic-reported time required for calculation and transcription of the data using cliexa-RA was 1 minute with an additional 10 second direct data integration to the EMR after form submission, as compared to the standard estimate of 5 minutes (personal communication by Robert Spencer, MD, 2018), a 77% reduction.

**Figure 2.**
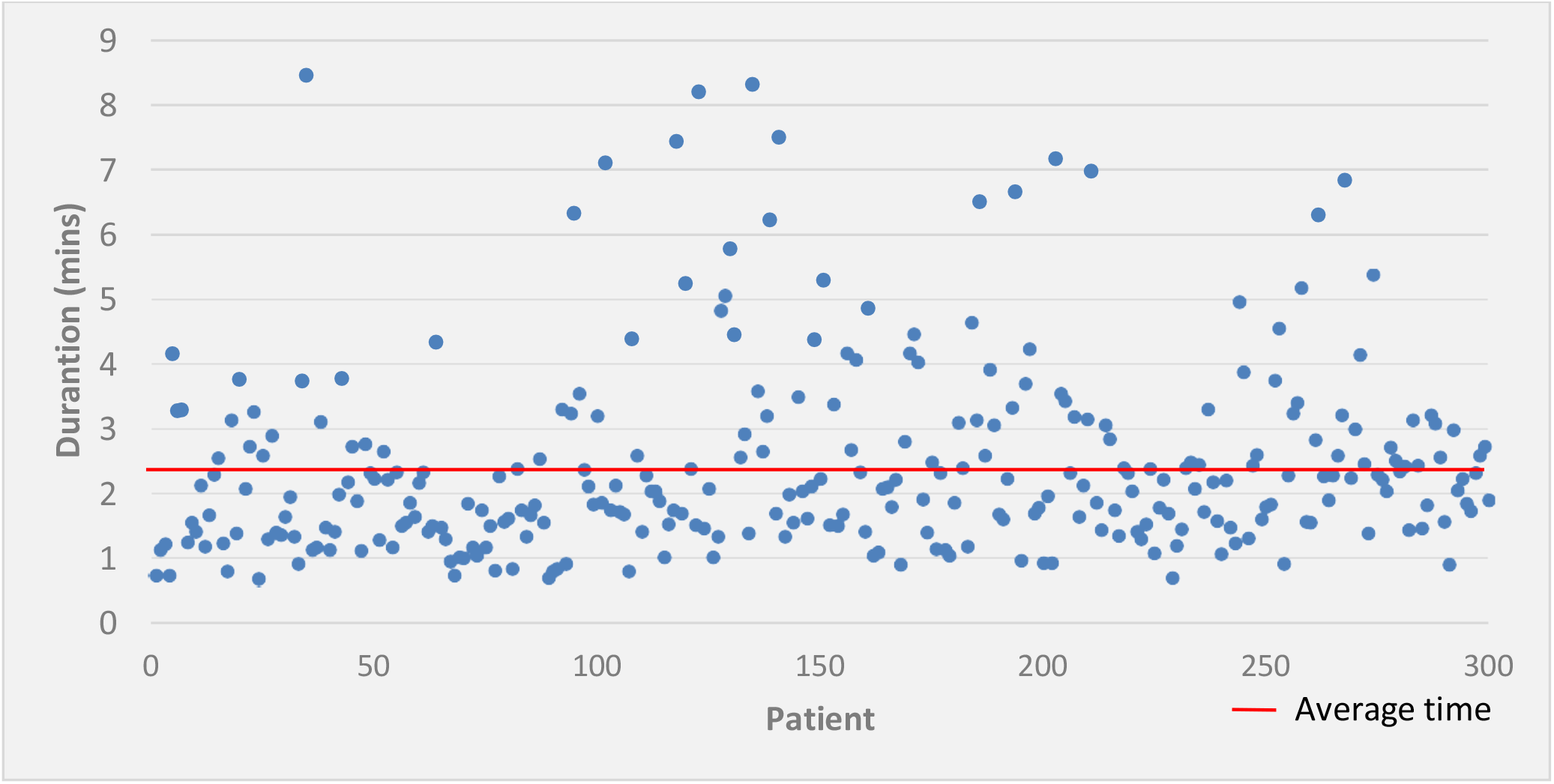
Time spent by patient to complete intake forms using cliexa-RA.

### Patient Experience

Based on survey results, 85% of patients said they would recommend cliexa forms to other clinicians. cliexa-RA scored an average of 3.57 out of 4 when compared to paper in ease of use. It scored an average of 3.61 in patient-reported reduction of time spent and 3.50 when patients are asked how easy the platform was to understand. Total average overall patient satisfaction with the cliexa-RA platform was scored at 3.55 out of 4 based on ease of use, time saving, preferred methods for clinical assessment and recommendations to other physicians (See Figure 3). Patients responded to the request for written recommendations for change with comments including “touch needs to work; more choices on answers; use paper; nothing, it’s perfect; room to explain some of answers; nothing, very easy to use.”

**Figure 3.**
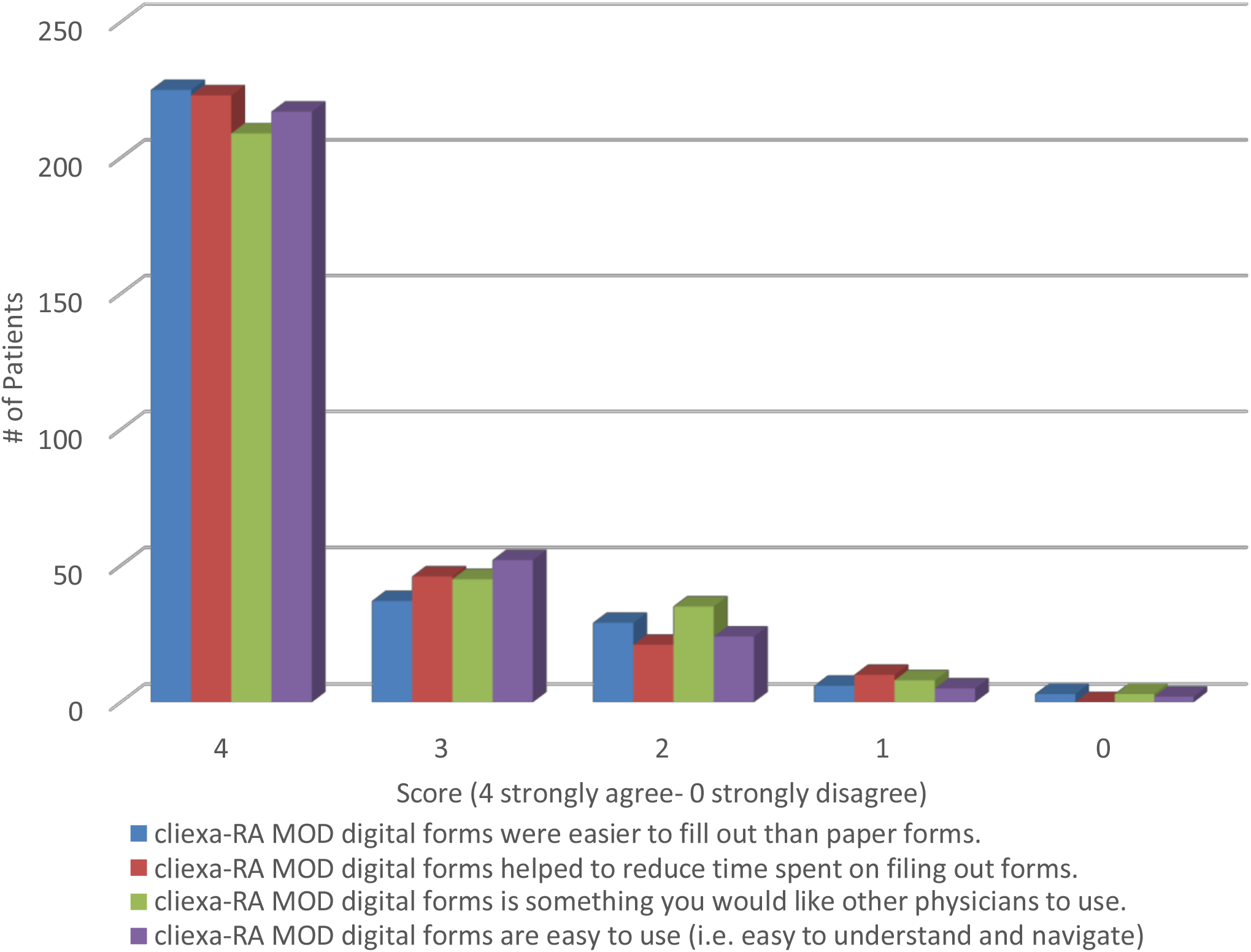
Patient engagement survey results.

### Physician Experience

Physician experience was measured through long-term engagement with cliexa-RA as well as testimonials. The pilot study led CAC to become cliexa’s customer with full integration pending in their Greenway Health EMR, and the implementation of pain and fibromyalgia assessment models in the existing platform. CAC physicians felt that the “cliexa platform makes electronic health records and documentation more efficient” (15).

The format and EMR upload capability ensure that this data is stored for future reference for each patient. Additionally, by providing a five-score reporting system, clinicians may integrate the data they have already collected using their score of preference to the new patient data outputs.

### Cost of Care and Population Health

Cost of care was measured by the reduction of clinic costs for annual MIPS reporting. To receive the largest possible reimbursement, a clinic must submit a full year of data to Medicare (16), costing on average $2,000 through a third-party reporting entity (personal communication by Robert Spencer, MD, 2018). This cost was reduced due to the automatic calculation and EMR compatible upload capabilities in the MIPS required format.

Data regarding immediate population health impacts were not available.

## Discussion

### Principal Results

Positive patient experience results in this study may be explained by reduced time, ease of use and the ability to transfer information from their intake form to their clinician without having to re-explain in the exam room. It was also reported that the digital format may have been easier for elderly patients with impaired vision to use due to the touch screen contrast and large font formatting (personal communication by Robert Spencer, MD, 2018). Additionally, RA patients typically have a high prevalence of comorbidities such as depression (33% in the U.S.), COPD (7.5% in the U.S.), elevated blood pressure (11.2% globally) and cardiovascular events (6.6% globally), (17). To address this need, the platform creates an opportunity for sharable quantified pain data for use in multiple disease states to better coordinate chronic disease management and increase patient health outcomes.

Patients have access on their personal devices through cliexa-RA mobile to additional disease activity measures, medication adherence and complication tracking, and medication reminders. With the application, the patient can report daily updates that are sent to the clinician. Custom alerts within the EMR or other communication methods such as emails or SMS are then available to care teams to set up additional follow up visits. The tracking tool allows patients to observe daily changes over time to identify patterns and improvements. Follow up updates can potentially reduce the incidence of ER visits and hospitalizations, thereby reducing healthcare costs and major disruptive events for the patient. This gives the patient the ongoing opportunity to advocate for their health.

Patient satisfaction is also heavily reliant on the professional satisfaction of their provider. In fact, patients of physicians with high levels of satisfaction were found to be happier with healthcare overall (18). However, in 2014, the Mayo Clinic published a study that found 54.4% of physicians to be experiencing at least one symptom of burnout, up from 45.5% in 2011 (19). Physician-patient interaction has changed significantly over time and Sinsky et al., note that 49.2% of a physician’s time is allocated to electronic health recording and desk work, and only 27% of the time is spent working directly with patients (5). These statistics can explain the aversion that some physicians feel to the implementation of process changes. With the traditional paper form process, a clinician spends valuable time calculating scores, reviewing patient data, and completing documentation for payors. cliexa-RA works to reduce the time spent by physicians on this process by creating quantified providing validated disease activity score outputs to enhance the efficiency and understanding of how the patient is doing as soon as they enter the exam room. CAC physicians highlighted the benefits of the data output from the platform addressing three of the nine required reporting points for MIPS (pain assessment, disease activity assessment, functional status) and disease prognosis, increasing plausible reimbursement and addressing billing rejection concerns.

Another consideration for adopting a digital platform is the customization for clinic needs and EMR integration, a process that often requires 6-9 months. Customization within the cliexa platform was completed on a one-day turnaround with development, staging and production updates.

The platform not only serves physician’s needs, but it also creates a valuable tool in the CMS managed care model for chronic disease. In this model, physicians are encouraged to remotely monitor patients to ensure ongoing care. Typical solutions to these changes are in the form of a one-size-fits-all bundled managed care platforms that can be costly and ill-fitting.

Cost of care can be measured both in fiscal savings and time output. Specialist clinics often carry heavy patient loads, limiting physician’s ability to calculate all five scores for each patient and therefore that information may not be collected. This can lead to audits, or compromised rates of reimbursement. With cliexa-RA, score output can be calculated and available for physicians with little time commitment from the patient or medical staff, potentially reducing clinic wait times and increasing physician face-time with patients. Cost savings can be estimated through the cliexa cost analysis tool which approximates the monetary value of staff time and the time saved daily by utilizing cliexa-RA. The Colorado Arthritis Center has five physicians, each seeing roughly 20 patients per day. With a standard paper process the medical staff spends approximately five minutes with each patient, or more than eight hours per day. When comparing this to the average hourly medical staff cost of $20, this process costs the clinic approximately $40,000 each year. With a 77% time savings for medical staff using the cliexa-RA platform, the CAC has the potential to save 30,800 each year in medical staff time savings alone^1^.

As a billable tool, physicians can remotely track patients and provide necessary care during critical treatment transition times through the use of a sister tool, cliexa-RA Mobile. Due to these capabilities, the cliexa-RA suite also has the potential to cut overall healthcare costs by reducing the risk of ER visits, which tends to lead to hospitalization if the patient is using injectable treatments.

Population health implications were not directly observable based on the data gathered, however there are numerous probable contributions that cliexa-RA could have. As a tool for gathering discrete data, cliexa-RA can be shared with other specialties, encouraging the managed care model for chronic disease. Collaboration among providers leads to a collaborative treatment effort that mimics a patient-centered home in which relevant data can be used to assess pain correlation (20). The steady collection of patient self-reported data incorporated into the patient’s electronic record is likely to improve disease management, leading to improved health outcomes (21) across all comorbidities and increasing self-efficacy. One study foundthat over a period of two years there was a significant correlation between the patient’s level of self-efficacy and their health status in relation to pain, mental health and general health. Additionally, this data will increase in value as a registry of population management and a data source for future RA research (21).

## Conclusion

Through the implementation of cliexa-RA in the patient intake, screening, and follow up processes, we found that patient and physician experience, and cost of care were all positively impacted. Opportunities exist for significant improvements in population health including the emphasis on the managed care model and reduction of overall healthcare costs.

## Data Availability

All data produced in the present work are contained in the manuscript

## Abbreviations

CAC: Colorado Arthritis Center
MIPS: Merit-based Incentive Payment System
RA: Rheumatoid Arthritis

## Acknowledgements

We thank Dr. Theodore Pincus for his contribution of the Rapid-3 assessment model used in cliexa-RA.

## Conflicts of Interest

Potential conflicts of interest include the contractual engagement during the development of the manuscript, and later, employment of author, Ashley Darnell, at cliexa, Inc. Additionally, private business interests with cliexa, Inc. exist for author, Robert Spencer, MD.

## Footnotes

1 Assuming eight hours per day, five days per week, 50 weeks per year.

## Notes

### Competing Interest Statement

The authors have declared no competing interest.

### Funding Statement

This study did not receive any funding

### Author Declarations

The study utilized de-identified, non-interventional data collected as part of routine clinical operations and workflow optimization. No protected health information (PHI) was collected or analyzed by the cliexa platform, as patient identifiers were excluded and replaced with internal identifiers for EMR linkage purposes. Based on these factors, the work is considered not human subjects research and therefore exempt from IRB review.

